# Learnings from the Australian First Few X Household Transmission Project for COVID-19

**DOI:** 10.1101/2022.01.23.22269031

**Authors:** Adrian J Marcato, Andrew J Black, James Walker, Dylan Morris, Niamh Meagher, David J Price, Jodie McVernon, the Australian FFX Household Transmission Project Group

## Abstract

**Background:** First Few “X” (FFX) studies provide a platform to collect the required epidemiological, clinical and virological data to help address emerging information needs about the COVID-19 pandemic.

**Methods:** We adapted the WHO FFX protocol for COVID-19 to understand severity and household transmission dynamics in the early stages of the pandemic in Australia. Implementation strategies were developed for participating sites; all household members provided baseline epidemiological data and were followed for 14 days from case identification. Household contacts completed symptom diaries and had respiratory swabs taken at baseline, day 7 and day 14, and day 28 where applicable. We modelled the spread of COVID-19 within households using a susceptible-exposed-infectious-recovered-type model, and calculated the household secondary attack rate and key epidemiological parameters.

**Findings:** 96 households with 101 cases and 286 household contacts were recruited into the study between April–October 2020. Forty household contacts tested positive for SARS-CoV-2 in the study follow-up period. Our model estimated the household secondary attack rate to be 15% (95% CI 8–25%), which scaled up with increasing household size. Children were less infectious than their adult counterparts but were also more susceptible to infection.

**Interpretation:** Our study provides important baseline data characterising the transmission of early SARS-CoV-2 strains from children and adults in Australia, against which properties of variants of concern can be benchmarked. We encountered many challenges with respect to logistics, ethics, governance and data management that may have led to biases in our study. Continued efforts to invest in preparedness research will help to test, refine and further develop Australian FFX study protocols in advance of future outbreaks.

**Funding:** Australian Government Department of Health

**Research in context:** *Evidence before this study:* The emergence of SARS-CoV-2 was initially characterised by uncertainty over key epidemiological, clinical and virological characteristics of the pathogen. We conducted a prospective household transmission study of confirmed cases of COVID-19 and their household contacts to collect data to understand severity and household transmission dynamics in Australia and add to the emerging evidence base for decision making. Large systematic reviews and meta-analyses of severity and transmission dynamics of SARS-CoV-2 in households have since been published, although estimates vary by setting.

*Added value of this study:* This is the first multi-jurisdictional prospective household transmission study of its kind for SARS-CoV-2 in Australia. Australia experienced low epidemic activity during the study period in 2020 due to robust public health and social measures including extensive PCR testing of symptomatic persons and isolation of all known contacts of confirmed cases. Hence, we describe the transmission dynamics in our cohort, i.e. in a low incidence setting and provide estimates of the household secondary attack rate, the relative susceptibility of children compared to adults, and transmission from children compared to adults.

*Implications of all the available evidence:* Our findings describe the epidemiology of COVID-19 in Australian households in 2020, and demonstrate the effectiveness of public health measures to limit transmission in this setting. Comparisons to other household transmission studies must be interpreted in light of the local epidemiology and context including study design, and sampling methods. Additional research is needed to incorporate genomic and serological data to further study transmission dynamics in our cohort. Continued development of the FFX study platform in Australia will enable integration into surveillance systems and help inform targetted public health responses to future infectious disease emergencies.

## Introduction

The global spread of the SARS-CoV-2 virus which causes coronavirus disease 2019 (COVID-19) was deemed a pandemic in March 2020.^1^ The emergence and global spread of SARS-CoV-2 was initially characterised by uncertainty over key epidemiological, clinical and virological characteristics of the pathogen, particularly, its ability to spread between humans and cause disease in a susceptible population.

The First Few “X” (FFX) study protocol for COVID-19 published by the World Health Organization (WHO) provides a platform to collect the required epidemiological, clinical and virological data to help address emerging information needs about the pandemic.^2-4^ The FFX study protocol is one of several protocols published by WHO as part of their UNITY study framework, which also includes standardised sero-epidemiological study protocols in household, health care and school settings amongst others.^5,6^

In February 2020, the eight Australian state and territory health departments together with the Commonwealth Department of Health and researchers from the Australian Partnership for Preparedness Research on Infectious Disease Emergencies (APPRISE) developed a national plan to implement the WHO FFX study protocol for COVID-19 in Australia.^7,8^ The Australian FFX Household transmission project aimed to inform understanding of local COVID-19 epidemiology in the early epidemic phases, and provide evidence for the development of guidelines and policy in specifically directing Australia’s ongoing public health response. The findings from this investigation are described here.

Australia’s first epidemic wave in 2020 was driven by returned international travellers and subsequent local transmission in major urban centres across the country. Public health and social measures were introduced to control the escalating epidemic, which included: border closures, expanded case management and contact tracing, and social measures such as density quotients in workplaces and public venues and lockdowns. Mandatory quarantine for returned international travellers was also introduced to reduce the risk of further importation. These measures drove incident cases in Australia to very low levels, and effective elimination (sustained periods of zero case incidence) was achieved in many states and territories by May 2020. A national strategy was set to pursue no community transmission of COVID-19 in the absence of widespread vaccine coverage.^9^

Breaches from the compulsory quarantine system for returned international travellers led to intermittent periods of local transmission in Australia, particularly in 2020 and the early stages of 2021. Australia’s second most populous state, Victoria, experienced a second epidemic wave of activity from late May 2020 to November 2020.

Several Australian states, including New South Wales (Australia’s most populous state), Victoria and the Australian Capital Territory now have established community transmission of SARS-CoV-2 due to the delta variant. As of December 13^th^ 2021, there have been 228,930 confirmed cases of COVID-19 in Australia, including 2104 deaths. Of these cases, 220,083 were locally acquired and the majority have been confirmed since June 2021.^10,11^

## Methods

### Study design, ascertainment and eligibility

We adapted the WHO UNITY FFX transmission study protocol for COVID-19, focusing on the household components, with a goal of recruiting 200 households into the project across participating sites.^3^ Participating sites included New South Wales (NSW; capital Sydney), Victoria (VIC; capital Melbourne), Western Australia (WA; capital Perth), South Australia (SA; capital Adelaide) and Queensland (QLD; capital Brisbane).

These adaptations included separating the study into two components: public health (data and viral swab collection as part of enhanced public health unit surveillance activities), and; additional research components (sequencing of positive samples and serology collection and analysis, not presented here), as detailed in Supplementary Table 1.

Laboratory confirmed index cases were recruited from the NSW, WA, and QLD state public health units where they were the first case identified in the household according to public health investigations and contact tracing. We recruited co-primary index cases where two household members tested positive within a 24-hour period and there was at least one other household member who was PCR-negative at baseline. In addition, we enriched for index paediatric cases by recruiting from the Royal Children’s Hospital Respiratory Infection Clinic (RCH) in VIC. Recruitment was active between April–October 2020, prior to the emergence of any variants of concern (Supplementary Figures 1–4).

Households were defined as two or more people living together in a domestic residence or a dwelling or group of dwellings with a shared space. Residential institutions were not included. All locally acquired cases were eligible for recruitment regardless of local source of infection provided they lived within an appropriate geographical area for logistics (i.e., metropolitan areas), and were not in mandated 14-day quarantine. All household members of eligible cases were required to provide their consent to participate. Hospitalised index cases were eligible for recruitment as we assumed that household contacts were exposed by the time hospitalisation of the index case has occurred. Households were excluded when all household members were infected at the time of the initial visit, making the direction of transmission events unclear and unobservable.

Epidemiological data were collected from confirmed cases and household contacts as close as possible to laboratory confirmation (day 0/baseline) of the index case, including health status interviews on days 7,14 and where available day 28. The questionnaires collected details on participant demographics, symptoms and vaccine and medical history (details provided in Supplementary Table 2). Household contacts also completed daily symptom diaries (via SMS) and provided specimens in line with Public Health Laboratory Network advice at baseline, days 7,14 and where available day 28. Respiratory swabs were professionally or self-collected depending on study site and were tested by polymerase chain reaction (PCR) in the state of collection.^12^ Households were cleared from the project at day 14 if all household contacts were symptom free and tested negative for COVID-19 at previous study timepoints (baseline and days 7/14). Index/primary cases did not complete symptom diaries or provide further swabs during their involvement in the study.

Deidentified data were collected and managed using REDCap electronic data capture tools hosted at The University of Melbourne. Ethics approval was not required for the FFX public health components being led through state and territory health departments as the project was recognised as an enhanced national public health surveillance activity. Ethics approval for the FFX project at the RCH site was obtained through the Murdoch Children’s Research Institute Ethics Committee (ref: 63666).

### Analysis

Descriptive analyses were performed to explore the characteristics of initially confirmed cases and their household contacts.

The household secondary attack rate (HSAR) was defined as the proportion of household contacts that were eventually infected in their study follow-up. We assumed that individuals tested positive for COVID-19 by PCR if and only if they had COVID-19 (i.e., the false positive rate is zero) and infected household contacts had at least one positive PCR test during their follow-up period. We classified all further detected cases within households as secondary cases and assumed that the primary case was the source of infection.

We characterised and modelled disease spread within households using an SEIR-type compartmental mathematical model previously developed for pandemic influenza^13-15^, and adapted it for COVID-19 according to early evidence about the incubation period and the generation interval.^16^ The model allows for pre- and asymptomatic infection status, and is age-structured allowing for age-specific contact rates.^17^ Adults were defined as 18 years old or older, and children were defined as less than 18 years old. The rate of transmission was allowed to scale depending on the household size. Model parameters were estimated using a bespoke Markov chain Monte Carlo method^15-16^; additional model details are outlined in the Supplementary Technical Appendix. Median posterior estimates and 95% credible intervals (CrI) are reported.

Statistical analysis was also conducted to support the choice of variables considered in the mathematical model, identify other variables that may be able to inform the mathematical model, and to align with global FFX and UNITY studies. We used logistic regression models to investigate the association between the HSAR and case- and household-level covariates. Multilevel mixed-effects logistic regression models were used to account for multiple observations per household in the contact-level HSAR analysis. The covariates used in these models are detailed in Supplementary Table 3. Households with co-primary cases were excluded from the statistical HSAR analysis but are included in the household model analysis.

Alpha was set to 0□05, and covariates that had a p-value of <0□2 in univariate regression analysis were included in the multivariable models for the different variable levels. Adjusted odds ratios, adjusted marginal estimates of the HSAR, and associated 95% confidence intervals (95% CIs) were produced for each included covariate.

Data cleaning and descriptive analyses were performed in R, (https://www.r-project.org/).^18^ Statistical HSAR analyses were performed in Stata version 16□0 (StataCorp LLC, College Station, Texas).^19^ All modelling and parameter estimation was performed using Julia 1.6.0 (https://julialang.org). ^20^

## Results

### Characteristics of FFX study population

We recruited 96 households with 101 confirmed index cases (due to co-primary cases) and 286 associated household contacts between April 2020 and October 2020. Three households had a false positive index case and were subsequently removed. Four households had incomplete study data. Supplementary Figure 1 shows recruitment into our study over time in relation to the number of locally acquired cases in Australia and in states that contributed data (Supplementary Figures 2–4).

FFX cases had a median age of 29 years (Interquartile range 15–42) and there were slightly more female cases than males. Thirty-five of the confirmed cases were children (<18 years old). Further case and contact participant characteristics can be seen in Table 1.The median household size was 4 (IQR 3–5) and ranged from 2–10 persons (Supplementary Figure 5).

**Table 1:** Characteristics of included case and household contact participants in the FFX project

|  | **Confirmed cases**<br>**(n = 101, from 96 households)** | **Household contacts**<br>**(n = 286)** |
| --- | --- | --- |
| ***Age, years*** |  |  |
| Mean (SD) | 28.0 (18.3) | 28.0 (19.3) |
| Median (IQR) | 29.0 (15.0–42.0) | 26.0 (11.0–44.0) |
| ***Age group, No. (%)*** |  |  |
| <12 | 21.0 (20.8) | 73.0 (25.5) |
| 12–17 | 14.0 (13.9) | 40.0 (14.0) |
| 18–49 | 55.0 (54.5) | 122.0 (42.7) |
| 50+ | 11.0 (10.9) | 51.0 (17.8) |
| ***Sex, No. (%)*** |  |  |
| Male | 48.0 (47.5) | 141.0 (49.5) |
| Female | 53.0 (52.5) | 144.0 (50.5) |
| Other | 0 (0) | 0 (0) |
| ***Received influenza vaccination in previous 12 months, No. (%)*** |  |  |
| Yes | 55.0 (54.5) | 131.0 (46.0) |
| No | 45.0 (44.6) | 151.0 (53.0) |
| Unknown | 1.0 (1.0) | 3.0 (1.0) |
| ***Ever had pneumococcal vaccine, No. (%)*** |  |  |
| Yes | 21.0 (20.8) | 82.0 (28.8) |
| No | 62.0 (61.4) | 145.0 (50.9) |
| Unknown | 18.0 (17.8) | 58.0 (20.4) |
| ***Pre-existing health conditions, No. (%)*** |  |  |
| Has pre-existing health conditions | 27.0 (26.7) | 87.0 (30.5) |
| Has no pre-existing health conditions | 74.0 (73.3) | 198.0 (69.5) |
| Asthma | 8.0 (7.9) | 32.0 (11.2) |
| Chronic respiratory condition (excluding asthma) | 1.0 (1.0) | 0 (0) |
| Cardiac disease | 1.0 (1.0) | 4.0 (1.4) |
| Immunosuppressive condition/therapy | 0 (0) | 1.0 (0.4) |
| Diabetes | 3.0 (3.0) | 8.0 (2.8) |
| Obese | 1.0 (1.0) | 5.0 (1.8) |
| Renal disease | 1.0 (1.0) | 1.0 (0.4) |
| Other condition(s) | 14.0 (13.9) | 41.0 (14.4) |

### Household transmission dynamics – mathematical modelling

Of the 286 household contacts recruited into the study, 40 tested positive for SARS-CoV-2 by PCR, with the majority (36/40, 90%) being detected and confirmed by the Day 7 timepoint. The modelling analysis is based on households with sufficient data (92 households comprising of 230 adults and 140 children). Of the included households, 68 had a single case only and experienced no secondary transmission. Final size distributions (i.e., the total number of individuals with laboratory-confirmed infections within a household over the period of monitoring) are shown in Figure 1.

**Figure 1:**
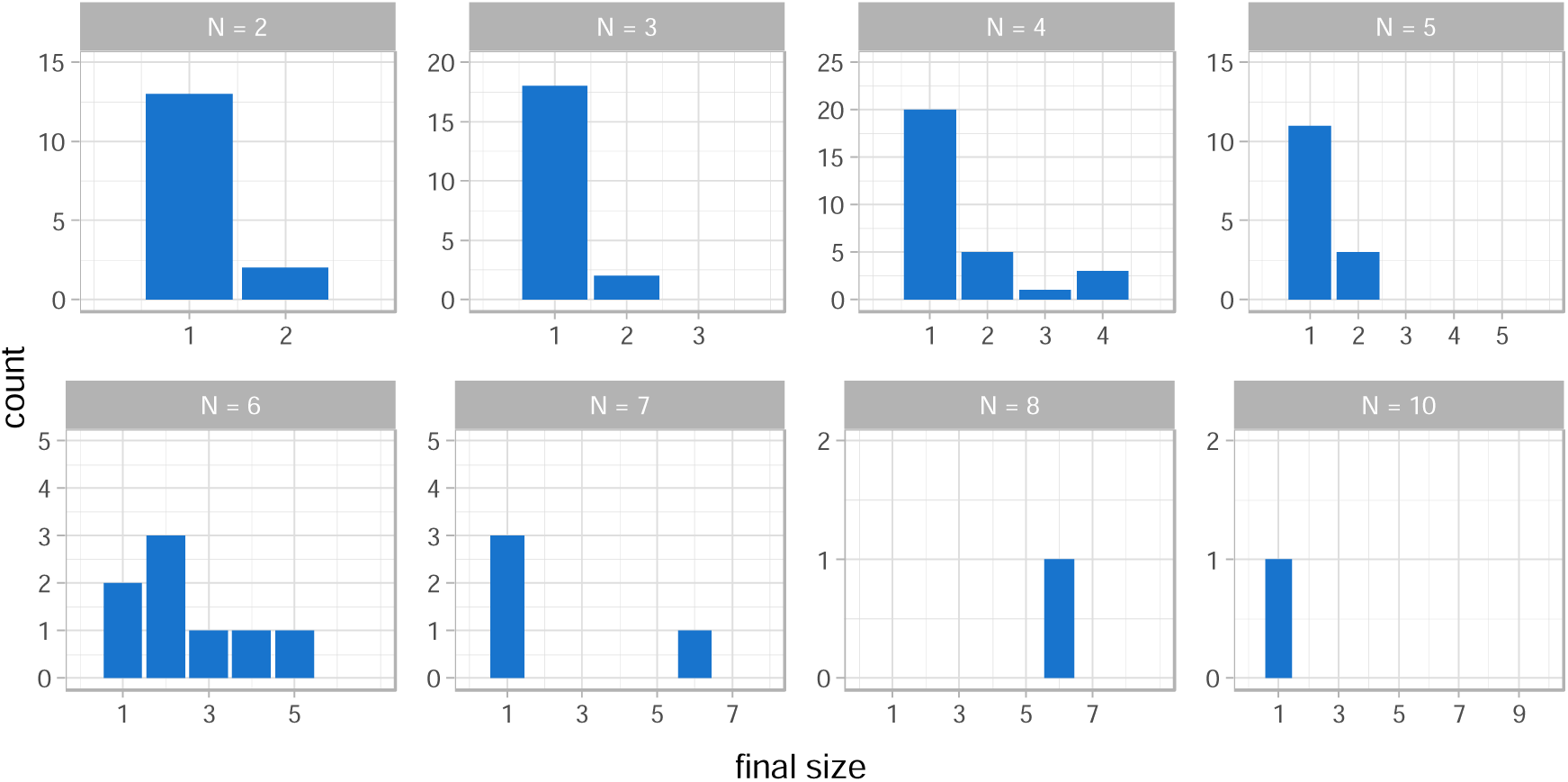
Final size distributions for the 92 households, where N is the size of the household. A final size of 1 indicates no secondary infections. There are no households of size 9 in the dataset.

Posterior distributions for the household secondary attack rate (HSAR) unstratified and stratified by household size, N (HSAR_N_), are shown in Figure 3. In both panels of this figure, HSAR is calculated as an average over the households in the dataset to account for the age-structured mixing and difference in adult-child transmissibility/susceptibility. The HSAR was estimated to be 15% (95%CrI 8–25%, Figure 2a) which increases with household size (Figure 2b).

**Figure 2:**
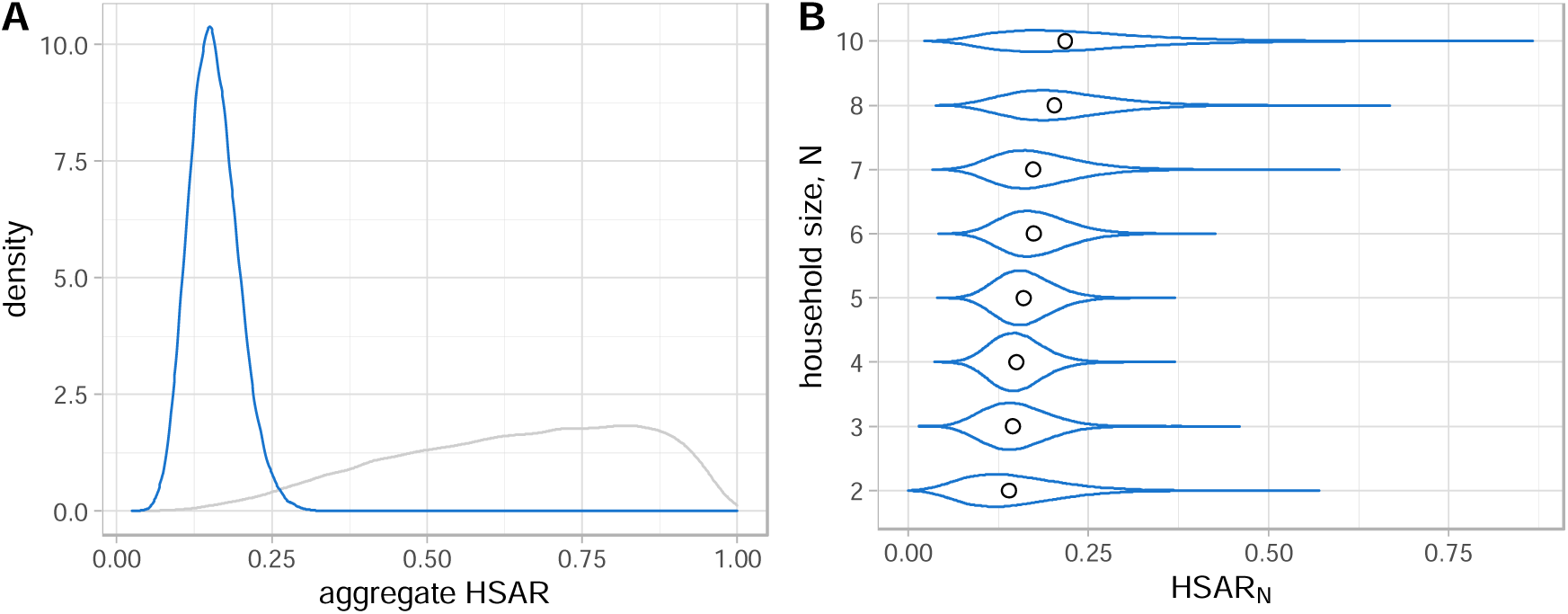
Posterior distributions for (A) the household secondary attack rate (HSAR) and (B) the household secondary attack rate conditional on household size N (HSAR_N_) shown in blue. The grey curve shows the prior distribution. In (B) the dots represent the median of the distributions. HSAR and HSAR_N_ are calculated as averages over the households in the study and over all ages.

**Figure 3:**
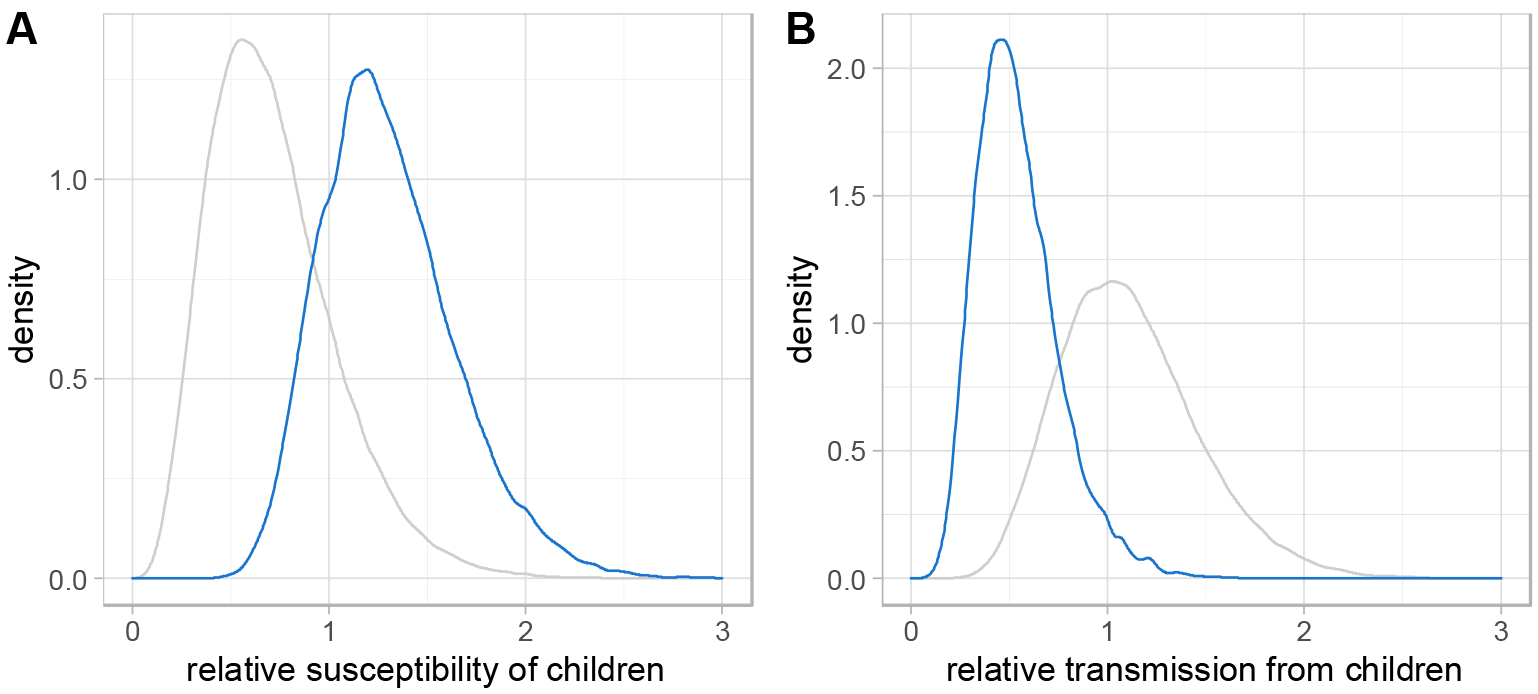
Posterior distributions (blue lines) for the relative susceptibility (A) and transmissibility (B) from children compared to adults, shown in blue. Prior distributions are shown in grey.

Adults had a higher likelihood of showing symptoms than children (Supplementary Figure 8). Children were found to be more susceptible than adults – the median posterior estimate of the relative susceptibility of children compared to adults was 1□26 (95%CrI 0.75–2.08) as seen in Figure 3A. Children were also less infectious than their adult counterparts – the median posterior estimate of relative transmissibility compared to adults was 0□52 (95%CrI 0.23–1.06), as seen in Figure 3B.

### Household transmission dynamics – statistical analysis

Using the contact-level mixed-effects logistic regression model and excluding households with co-primary cases, the HSAR estimate was found to be 12% (95%CI 7–17%). Details of the multivariable logistic regression models at the various factor-levels are presented in Supplementary Table 4. The odds ratio estimate for household size was 1□31 (95%CI 0.97–1□77, p=0.080), representing an average 31% increase in the odds of secondary infection within the household for each one person increase in household size. There is some evidence to suggest that HSAR is associated with the relationship between cases and their contacts – parents/guardians/carers and siblings had lower odds of being a secondary case when children were the primary case. The other covariates included in the multivariable models were not found to be associated with the HSAR.

### Severity

Four confirmed cases were hospitalised during their follow-up period (Case hospitalisation rate, 2.8% (4/141), 95%CI 0.9–7.5%) and no deaths were reported in our cohort.

Overall, 31.9% (45/141) of confirmed cases were asymptomatic (95%CI 24–40%). 10/101 (9.9%) were asymptomatic primary cases at baseline and 35/40 (87.5%) secondary cases were asymptomatic during their follow-up. Symptoms experienced by household contacts by COVID-19 status can be seen in Supplementary Figure 6.

## Discussion

Our household transmission study estimates the HSAR in Australia to be 15% (95%CrI 8–25%) prior to the emergence of variants of concern. We demonstrate that the HSAR increases with household size. Children were relatively more susceptible to infection compared to adults when exposed and were also less infectious than their adult counterparts.

The ‘gold-standard’ mathematical model captures the complex timing and dynamics of transmission in households. Thus, we believe these results to be more robust than those produced by the statistical models. Associations in the statistical modelling need to be taken with caution due to the small sample size and our underlying assumption that all cases we observe in our households are attributed to the primary case – an assumption that is not required in the mathematical model. However, the statistical model results are important as they are broadly consistent with the results from the robust mathematical modelling approach, and represent the standard analytic method that is used to analyse such household transmission studies. They are presented here such that results from our cohort may be fairly compared to other international studies.

Our HSAR estimate is consistent with estimates from two systematic review and meta-analyses of household transmission.^21,22^ We note that our results differ from similar household transmission studies including studies based on the WHO UNITY protocols, such as the FFX study conducted in the UK, which estimated a higher base HSAR that decreased with increasing household size.^23-33^ Other studies using population surveillance data, which represent transmission within a broader range of settings than the household, have estimated lower relative susceptibility to SARS-CoV-2 infection for children compared to adults.^34,35^

It can be difficult to make direct comparisons between studies that are conducted in different countries and settings due to the unique features of local epidemics and adaptations required for implementation. Studies should be interpreted in light of the local epidemiology and context – considerations should be made for the surveillance and contact tracing capacity, local incidence of COVID-19 cases during study implementation, predominant circulating SARS-CoV-2 variant, the timing and duration of the study, and study design including case ascertainment strategies and specimen sampling methods. Characteristics of individuals affected by COVID-19 and recruited into the study such as socioeconomic status, occupation and size of recruited households may also be significantly different across these studies, and therefore may influence aggregate outcomes. Additionally, differences in public health interventions such as: test, trace and isolate capacity and practices; behavioural and distancing measures; mobility restrictions; communication campaigns; and varying degrees of community engagement and cohesion in response, could also help to explain how estimates may vary across countries and settings.

We note ascertainment and recruitment bias in our study cohort that may contribute to some of the differences we observe to other studies – we excluded households where all members of these households were already infected at baseline. This was more likely to exclude smaller households than larger households for participation, and subsequently may have resulted in the HSAR for smaller households being underestimated. Our modelling outputs are therefore influenced more strongly by larger households, particularly three large outbreaks in households with more than five household members. These may be outliers and as such the observed effect could disappear if more data had been collected including from smaller households who experienced rapid transmission making them ineligible for recruitment. Additional sources of data could help us understand the extent to which our results are influenced by our inherent study biases and if our HSAR estimate is an underestimate, or if it is rather a feature of Australia’s unique epidemiology, i.e. transmission in a low incidence setting with stringent public health and social measures to reduce within-household and community transmission.

We did not observe longer chains of infection in households that had detected secondary transmission. As a result, there were insufficient data to confidently estimate other quantities of interest such as the incubation period, and the pre-symptomatic and symptomatic infectious periods. Although some households experienced larger absolute numbers of cases, in the majority of such households most individuals were already infected at the recruitment baseline or initial swabbing time point (90% of secondary cases were positive by day 7 testing). These outcomes were expected especially as public health units provided extensive advice to reduce the probability of additional spread within the household, including advice on mask use, and how to isolate from each other in their homes. Whilst not the case in this cohort, some cases were removed from their household to further mitigate the risk of spread if their home environment was not suitable for quarantine.

We conducted sensitivity analyses to consider how the arbitrary age cut-off of 18 years to define adults and children and the use of our contact matrices were impacting our results. We explored age cut-offs of 8,13 and 16 years of age. We found that the estimated HSAR was not sensitive to changes in the age cut-off (Supplementary Figure 8). There are small differences in the probability of symptom onset for the different age cut-offs (Supplementary Figure 9), although these appear to be centred on the same values. The age cut-off of 16 yielded posterior estimates for the probability of symptom onset that were very similar. There was no sensitivity to the contact matrix being used – this is likely a result of the large number of households who experienced no secondary transmission.

Our study has several strengths: This is the first multi-jurisdictional household transmission study of its kind for SARS-CoV-2 in Australia. We provide insights into household transmission with testing of known household contacts regardless of symptoms in a sustained low incidence setting, where there is more certainty about the source of SARS-CoV-2 transmission being from within the household, rather than the community, compared to a higher incidence setting. The pre-existing relationship between public health departments and APPRISE researchers was an enabling factor to provide capacity for the implementation of the study, as Australian health departments were prioritising hospital preparedness and scaling up testing and contact tracing in early 2020 when this study commenced. Our study enriched for paediatric cases through recruitment at the RCH site – children were generally not index cases at the other sites, and as such this recruitment strategy provided us with unique insights into household transmission from children in the Australian context.

Operationally, our data fields were aligned with the National Notifiable Diseases Surveillance Scheme to harmonise with enhanced surveillance efforts and reduce duplication of data collection where possible. Our bespoke REDCap database provided a central repository to analyse FFX data as a national dataset. Analysis and reporting of FFX data was performed in real time to key national and international stakeholders including, the Communicable Diseases Network Australia (CDNA), WHO Headquarters and the WHO Western Pacific Regional Office.

The lack of an Australian specific protocol with a pre-determined implementation strategy led to issues with logistics, and made it difficult to obtain the relevant ethics and governance approvals for all associated research components. We originally anticipated a 6–8 week window of intense recruitment in line with a short and sharp epidemic in early 2020. Strong social and public health control measures including border closures and mandated hotel quarantine reduced case numbers and subsequently the number of eligible cases and households. Two of our sites (QLD and SA) had sustained zero community transmission of COVID-19 by the time they were ready to recruit in April 2020 and WA achieved this in May 2020 after only recruiting four households.

We were able to recruit more as epidemic activity increased in VIC and NSW in mid-2020, but case ascertainment in Victoria was limited due to recruitment being limited to the paediatric hospital site. These factors prolonged the duration of our study and may have further contributed to our ascertainment bias.

Future research will also involve further collection and analysis of associated genomic and serological data in the FFX research components to better understand and confirm the transmission dynamics in our cohort. Genomic data can help confirm our classification of individuals as we assumed additional cases in the household were attributed to the index case. Serological data may identify historic infections in individuals who continue to present as PCR positive but are non-infectious. Serological data may also be important to identify previously undetected infections in household members especially as the rate of false negatives from PCR may not be insignificant.^36^ Together these can provide more accurate data to classify household members and subsequently inform attack rate calculations.

Our study provides important baseline data characterising the transmission of early SARS-CoV-2 strains from children and adults in the Australian context, against which properties of emerging variants of concern such as the Alpha and Delta strains can be benchmarked. ^37-40^ We plan to follow our recruited FFX households longitudinally to continue to develop our understanding of household transmission and immunity in the context of emerging variants of concern. This study will be conducted as Australia’s vaccination program continues and throughout the eventual establishment of community transmission of SARS-CoV-2 in Australia. Research is also currently underway to formally evaluate the implementation of our FFX study to help consolidate on lessons learnt and inform preparedness efforts for future FFX studies in Australia for COVID-19 or other infectious disease emergencies.

## Conclusion

The Australian FFX project for COVID-19 has been useful to provide valuable insight into the epidemiology of SARS-CoV-2 in Australia despite encountering many challenges in the planning and implementation phases with respect to logistics, ethics, governance and data management. Continued efforts to invest in preparedness research will help to test, refine and further develop Australian FFX study protocols in advance of future outbreaks of concern and ensure they are embedded in pandemic response plans.^41,42^ Being able to rapidly activate and provide high-quality information in real-time will be useful for epidemic situational assessment and modelling studies in response to future outbreaks of concern, to ensure a more proportionate, equitable and targeted public health response and help reduce disease impact.

## Data Availability

Data from the Australian FFX Project is subject to approval by the FFX Project Data Governance Committee. Please contact the authors for additional information.

## Acknowledgements

The Australian FFX Household Transmission Project Group (alphabetical order): Ross Andrews, Laura Bannerman, Christina Bareja, Andrew Black, Douglas Boyle, Georgina Collins, Jo Collins, Loral Courtney, Nigel Crawford, Katina D’Onise, Lucy Deng, Kate Dohle, Andrew Dunn, Paul Effler, James Fielding, Erin Flynn, Rob Hall, Sonia Harmen, Troy Laidlow, Eileen Lam, Adrian Marcato, Jodie McVernon, Niamh Meagher, Adriana Milazzo, Caroline Miller, Dylan Morris, Janette Mulvey, Sera Ngeh, Genevieve O’Neill, Kate Pennington, Priyanka Pillai, Ben Polkinghorne, David Price, Victoria Pye, Joshua Ross, Freya Shearer, Miranda Smith, Paula Spokes, Andrew Steer, Mark Taylor, Shidan Tosif, Florian Vogt, James Walker, Nicholas Wood.

The Communicable Diseases Network Australia, for their input in the planning stages of the project, oversight and review of the article.

## Supplementary Appendix

**Supplementary Table 1:** Australian FFX Household Transmission Project Components and details

|  | Summary | Funding source |
| --- | --- | --- |
| *Public health component* | - Data collection from confirmed cases as close as possible to laboratory confirmation including status interviews on days 7,14 ± 28<br>- Data collection from household contacts including daily symptom diaries and status interviews on days 7,14, ± 28<br>- Specimens at days 0,7,14 ± 28 from household contacts. Professionally collected or self-collected depending on site, in line with the Australian Public Health Laboratory Network advice<br>- Households to end participation where the entire household is symptom and COVID-19 free at the day 14 time point | Australian Commonwealth Department of Health |
| *Research component 1 – components of protocol not deemed to be essential public health activity in February 2020* | - Sequencing of previously collected positive specimens in the public health project<br>- Collection of blood sample from household contacts after any self-isolation/quarantine periods have been served | APPRISE Centre of Research Excellence |
| *Research component 2 – extended follow-up of FFX cohort over 3 year period* | - Repeat serology from current recruits<br>- Ongoing FFX recruitment – trialling FluTracking arm and Aboriginal Community Controlled Health Organisation led First Nations pilot study | Australian National Health and Medical Research Council (partnership grant in collaboration with the Australian Commonwealth Department of Health and Hunter New England Local Health District) |

**Supplementary Table 2:** Data collected in the Australian FFX questionnaires

| Case data | Contact data |
| --- | --- |
| - Demographic data; age, sex, pregnancy<br>- Aboriginal and Torres Strait Islander status<br>- Household demographics; household size and number of bedrooms, postcode<br>- Symptom data (including symptom diaries where relevant)<br>- Comorbidity data<br>- Laboratory data (swabs)<br>- Influenza/pneumococcal vaccination data<br>- Follow up data; including hospitalisation status at study time points | - Demographic data; age, sex, pregnancy<br>- Aboriginal and Torres Strait Islander status<br>- Relationship to case, and extent of contact<br>- Symptom data (including symptom diaries)<br>- Comorbidity data<br>- Laboratory data (swabs)<br>- Influenza/pneumococcal vaccination data<br>- Follow up data; including hospitalisation status at study time points |

**Supplementary Table 3:**
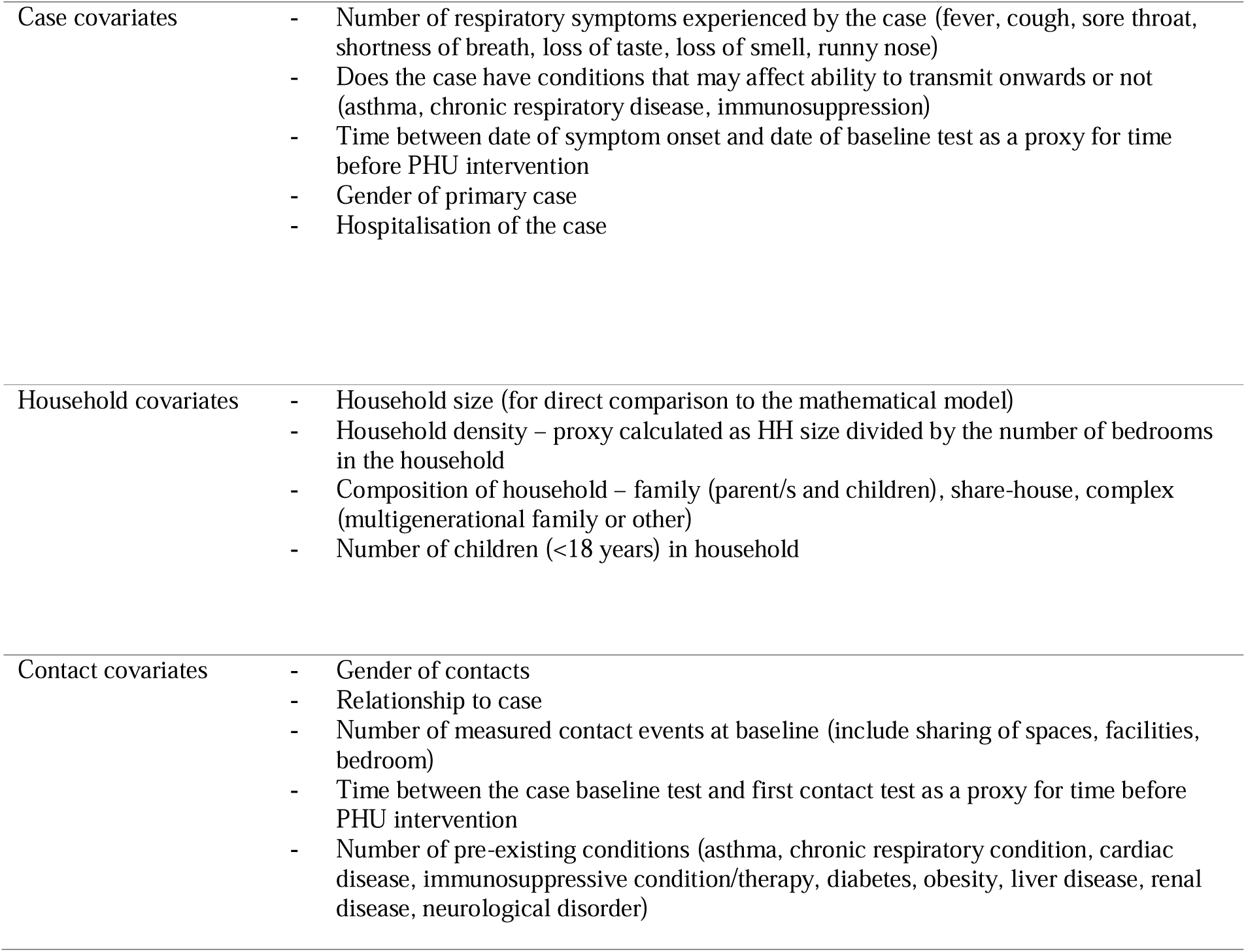
Covariates explored in the univariate and multivariable logistic and negative binomial regression models

|  |  |
| --- | --- |
| Case covariates | - Number of respiratory symptoms experienced by the case (fever, cough, sore throat, shortness of breath, loss of taste, loss of smell, runny nose)<br>- Does the case have conditions that may affect ability to transmit onwards or not (asthma, chronic respiratory disease, immunosuppression)<br>- Time between date of symptom onset and date of baseline test as a proxy for time before PHU intervention<br>- Gender of primary case<br>- Hospitalisation of the case |
| Household covariates | - Household size (for direct comparison to the mathematical model)<br>- Household density – proxy calculated as HH size divided by the number of bedrooms in the household<br>- Composition of household – family (parent/s and children), share-house, complex (multigenerational family or other)<br>- Number of children (<18 years) in household |
| Contact covariates | - Gender of contacts<br>- Relationship to case<br>- Number of measured contact events at baseline (include sharing of spaces, facilities, bedroom)<br>- Time between the case baseline test and first contact test as a proxy for time before PHU intervention<br>- Number of pre-existing conditions (asthma, chronic respiratory condition, cardiac disease, immunosuppressive condition/therapy, diabetes, obesity, liver disease, renal disease, neurological disorder) |

**Supplementary Table 4:** Results from the multivariable logistic regression models of HSAR. Covariates were included in the multivariable logistic regression models if they had a p-value of <0.2 in univariate regression analysis. The estimates presented here are exclusive of households with co-primary cases.

| Covariate | Variable level | Adjusted odds ratio (95% CI) | p-value | Adjusted HSAR estimate (95% CI) |
| --- | --- | --- | --- | --- |
| *Household-level model (n=91)* |  |  |  |  |
| *Household size<sup>#</sup>* | 2 | Overall – 1.35<br>(0.99, 1.84)* | 0.054* | 0.11 (0.02, 0.21)* |
|  | 3 |  |  | 0.15 (0.07, 0.24)* |
|  | 4 |  |  | 0.20 (0.11, 0.28)* |
|  | 5 |  |  | 0.26 (0.15, 0.35)* |
|  | 6 |  |  | 0.33 (0.16, 0.46)* |
|  | 7 |  |  | 0.41 (0.16, 0.60)* |
|  | 8 |  |  | 0.49 (0.15, 0.75)* |
|  | 9 |  |  | 0.57 (0.15, 0.90)* |
|  | 10 |  |  | 0.65 (0.24, 0.96)* |
| *Case-level model (n=91)* |  |  |  |  |
| *Ever hospitalised (from case-level model)* | No | Ref |  | 0.21 (0.13, 0.30) |
|  | Yes | 2.56 (0.29, 22.34) | 0.395 | 0.40 (0, 0.90) |
| *Number of transmitting conditions (asthma, chronic respiratory disease, immunosuppression)* | 0 | Ref |  | 0.20 (0.11, 0.29) |
|  | 1 | 2.75 (0.61, 12.34) | 0.186 | 0.40 (0.07, 0.74) |
| *Contact-level model (n=276)* |  |  |  |  |
| *Multilevel mix effects logistic regression model, incorporating clustering by household* |  |  |  |  |
| *Contact relationship to case* | Child | Ref | - | 0.22 (0.10, 0.33) |
|  | Other*** | 0.60 (0.01, 4.14) | 0.271 | 0.08 (0, 0.23) |
|  | Parent/guardian/carer | 0.07 (0.01, 0.39) | <0.01 | 0.04 (0, 0.09) |
|  | Partner/spouse | 0.57 (0.15, 2.24) | 0.422 | 0.16 (0.06, 0.27) |
|  | Sibling | 0.23 (0.04, 1.16) | 0.074 | 0.1 (0.01, 0.17) |
| *Sex* | Female | Ref | - | 0.10 (0.04, 0.14) |
|  | Male | 2.37 (0.80, 6.95) | 0.117 | 0.14 (0.08, 0.20) |
| *Number of pre-existing conditions (as defined in Supplementary Table 3)* | 0 | Ref | - | 0.10 (0.05, 0.14) |
|  | 1 | 6.37 (1.40, 28.89) | 0.016 | 0.26 (0.10, 0.39) |
|  | 2 | 2.47 (0.10, 62.85) | 0.585 | 0.16 (0, 0.42) |

\* Results from the univariate regression models are presented where only one covariate was eligible to be
included in the multivariable model
\*\*\* Includes grandparents, grandchildren, partners of household contacts, housemates

**Supplementary Figure 1:**
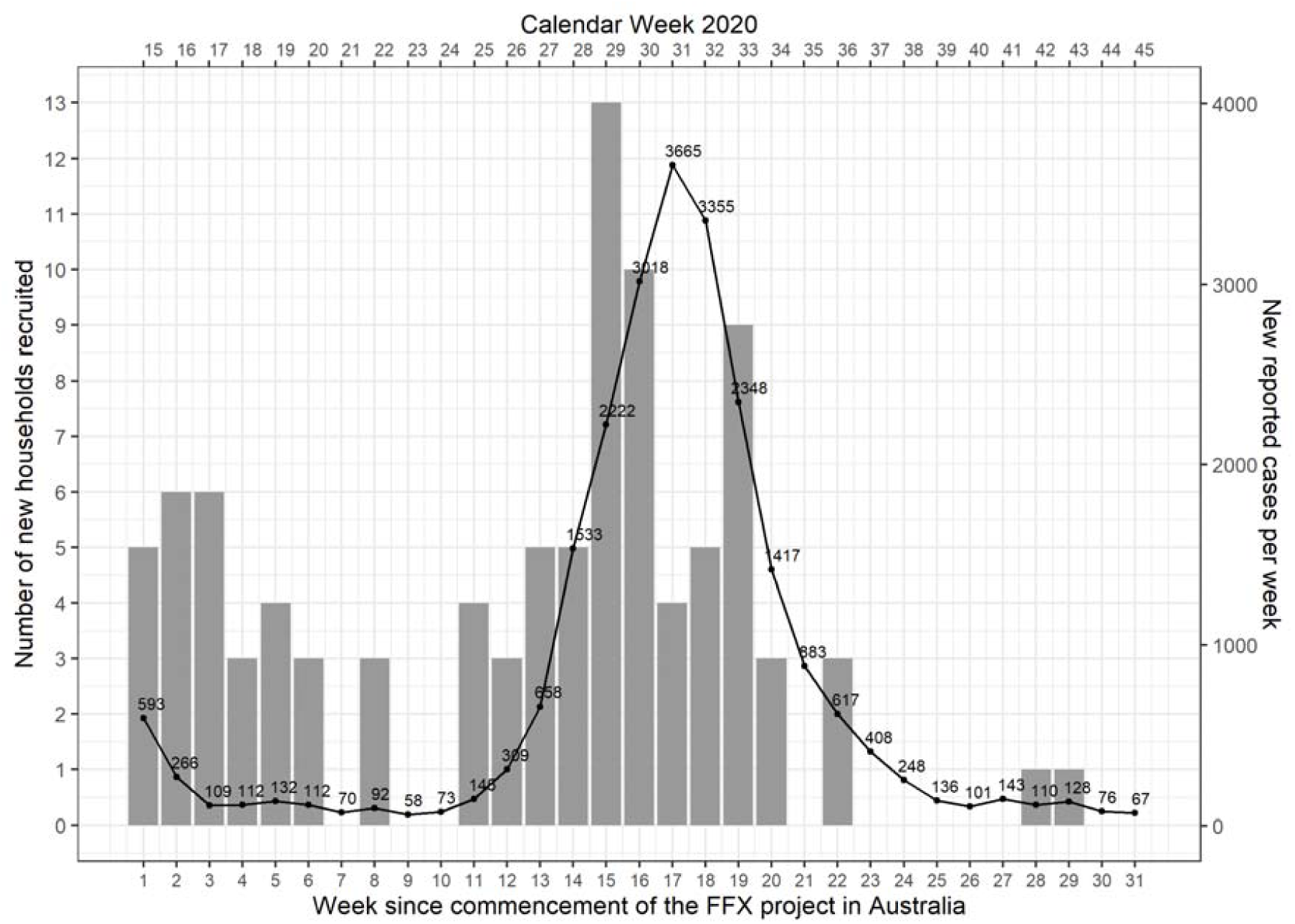
Recruitment into the Australian FFX project (bars) and confirmed cases across Australia (line). Note that the epidemic curve represents includes all cases reported in Australia from the commencement of the project (April 6^th^ 2020), and not necessarily all eligible local cases for inclusion into the project.

**Supplementary Figure 2:**
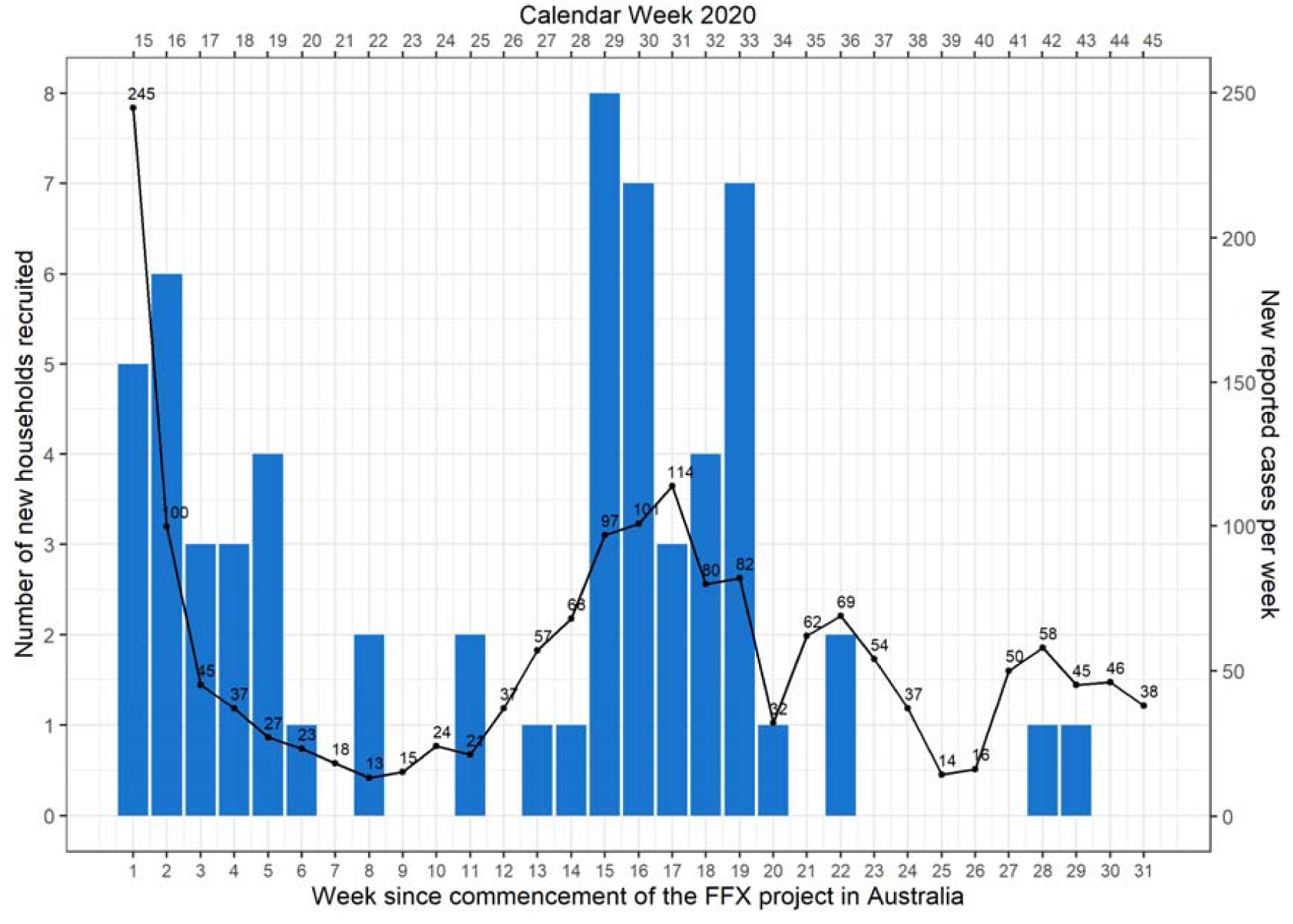
Recruitment into the Australian FFX project (bars) and confirmed cases in New South Wales (line). Note that the epidemic curve represents includes all cases reported in New South Wales from the commencement of the project (April 6^th^ 2020), and not necessarily all eligible local cases for inclusion into the project.

**Supplementary Figure 3:**
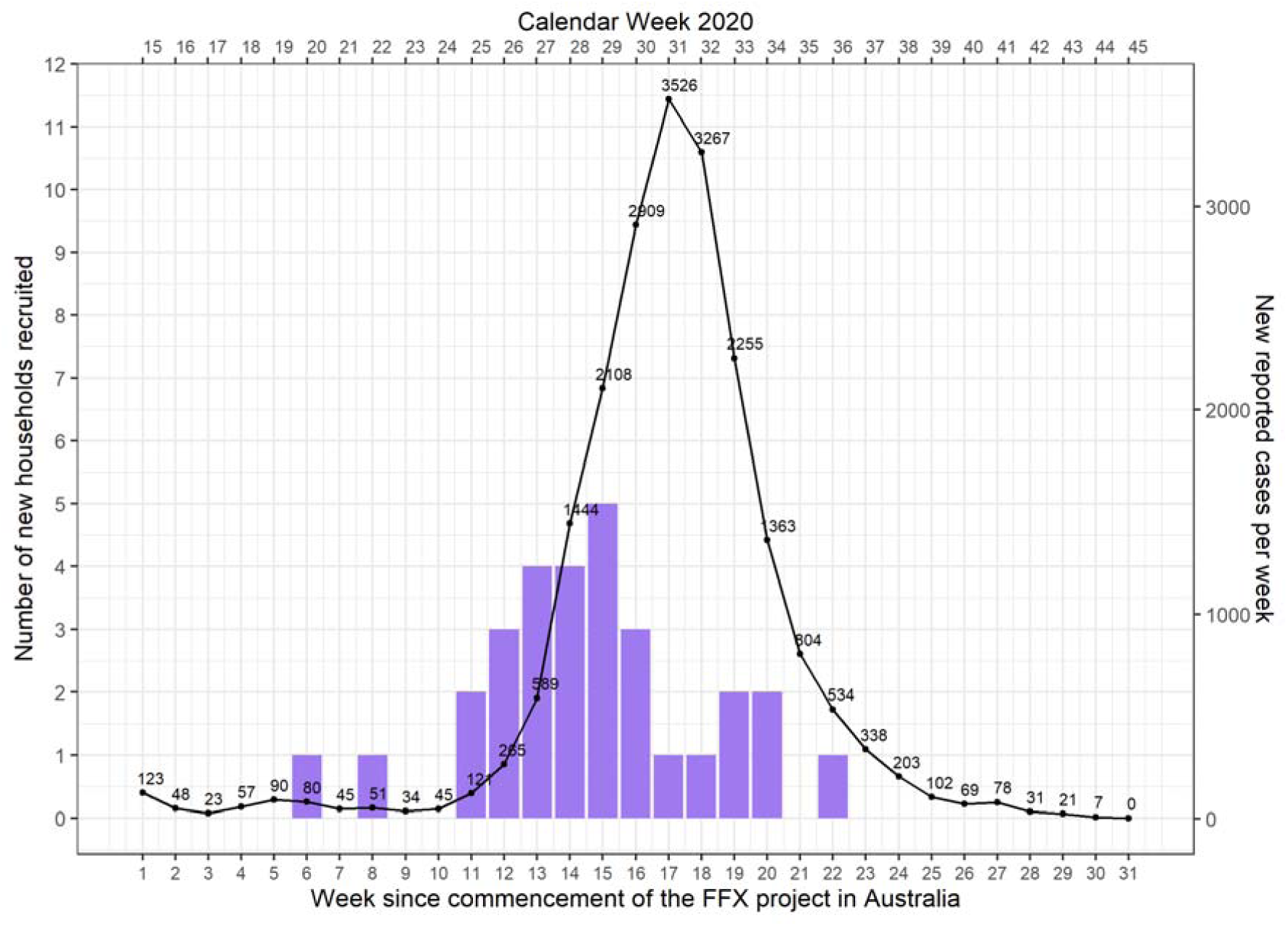
Recruitment into the Australian FFX project (bars) and confirmed cases in Victoria (line). Note that the epidemic curve represents includes all cases reported in Victoria from the commencement of the project (April 6^th^ 2020), and not necessarily all eligible local cases for inclusion into the project identified at the Royal Children’s Hospital.

**Supplementary Figure 4:**
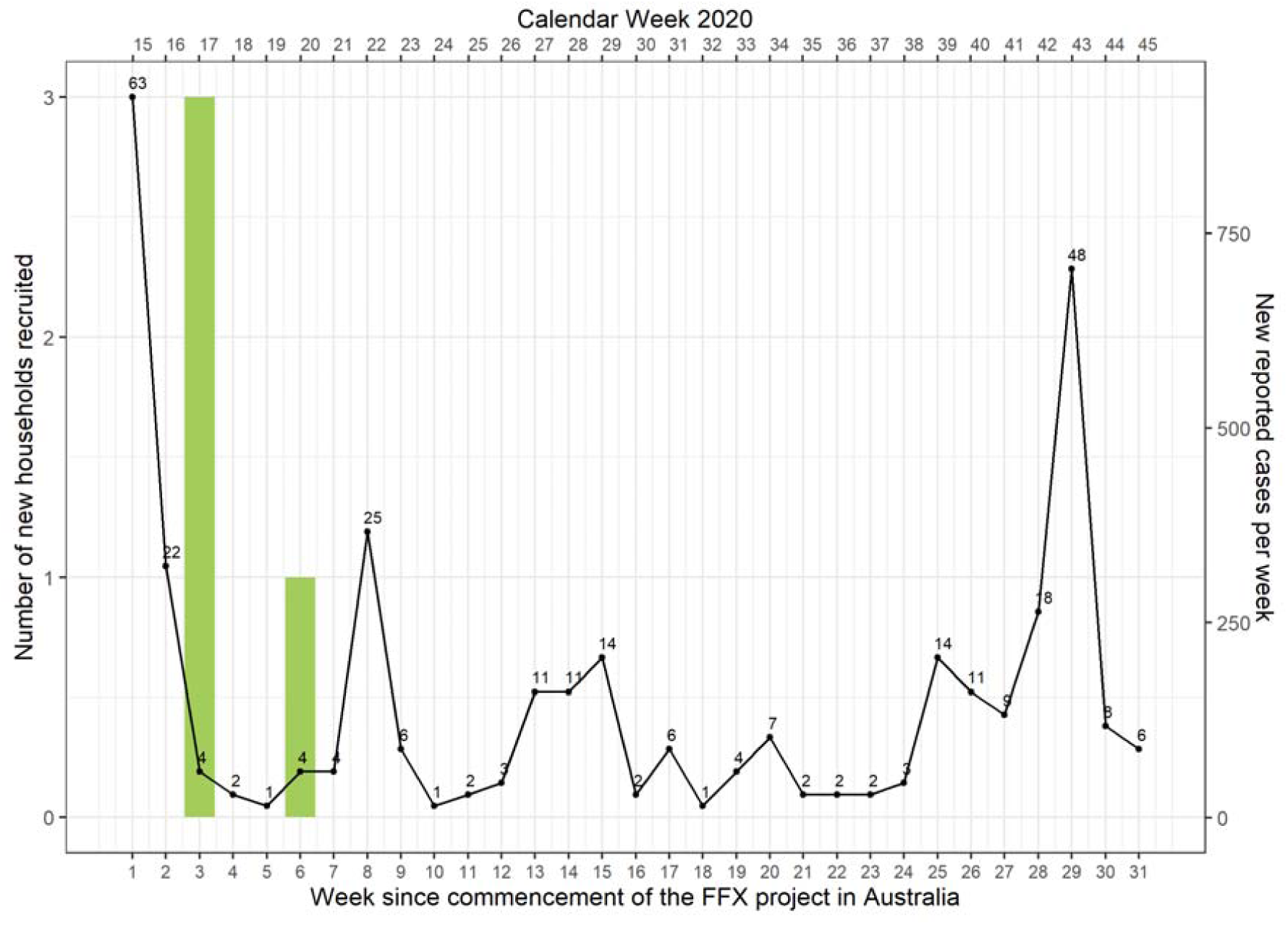
Recruitment into the Australian FFX project (bars) and confirmed cases at the Western Australia (line). Note that the epidemic curve represents includes all cases reported in Western Australia from the commencement of the project (April 6^th^ 2020), and not necessarily all eligible local cases for inclusion into the project.

**Supplementary Figure 5:**
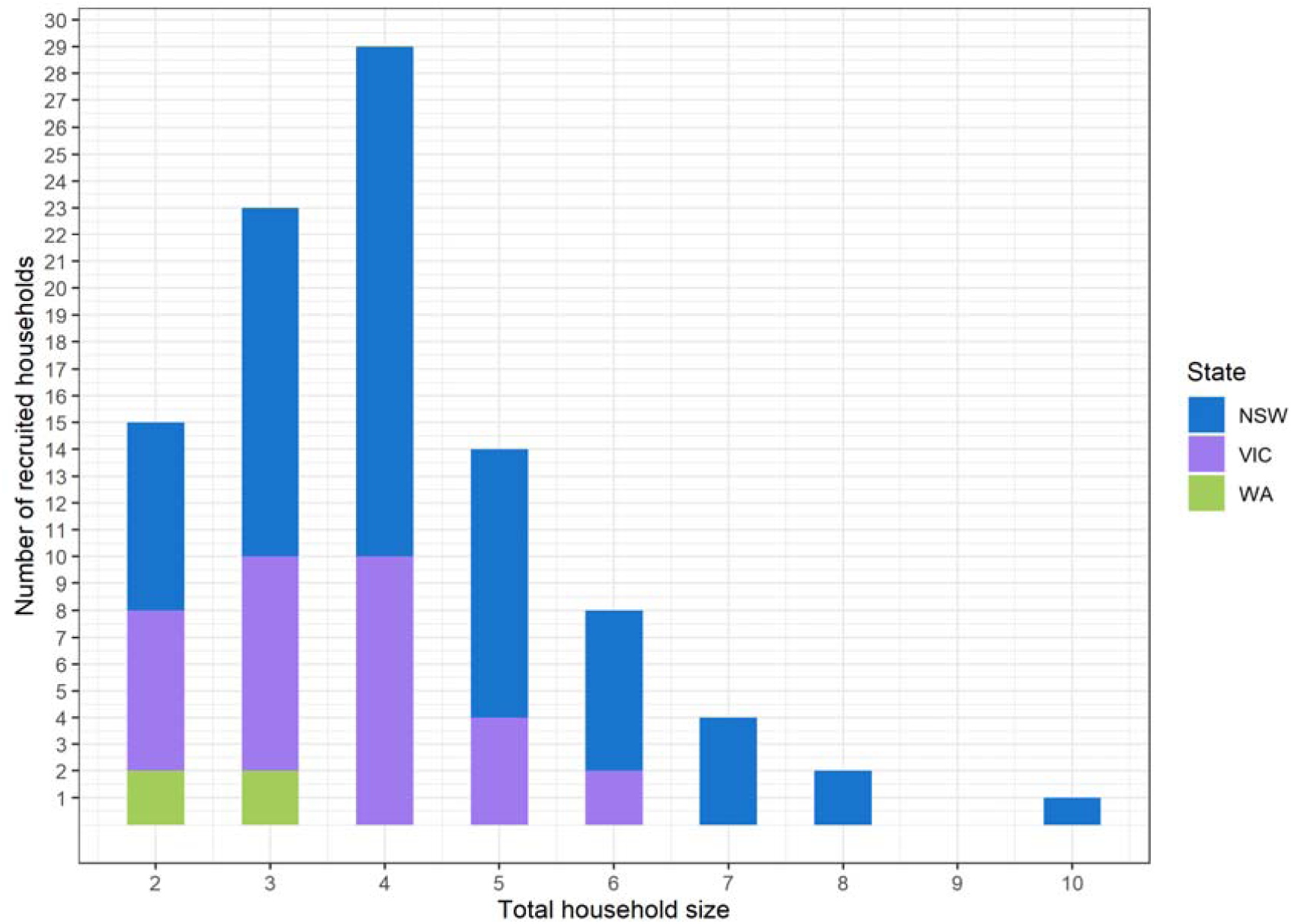
Histogram of recruited household sizes by state.

**Supplementary Figure 6:**
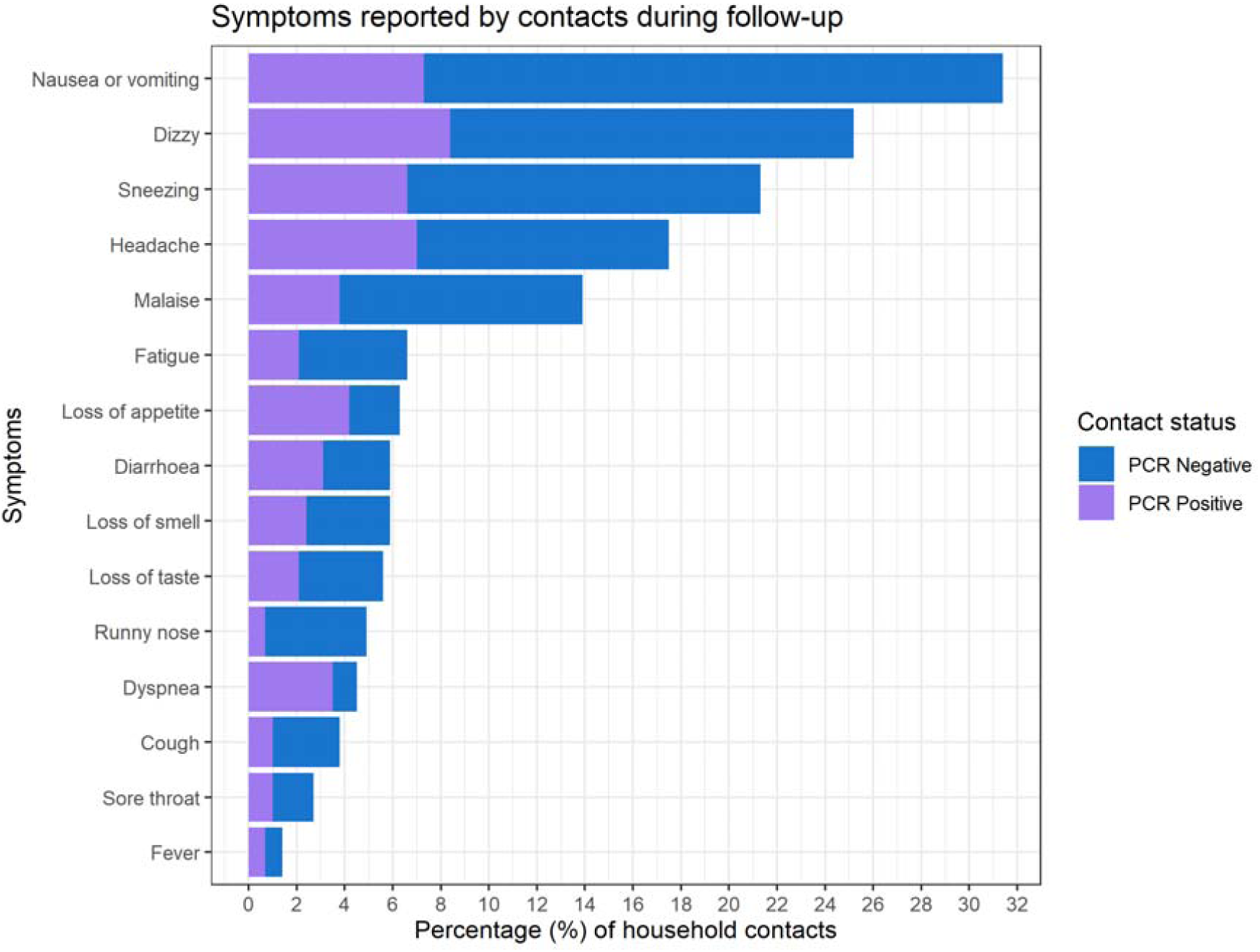
Symptoms reported by household contacts during study follow-up.

**Supplementary Figure 7:**
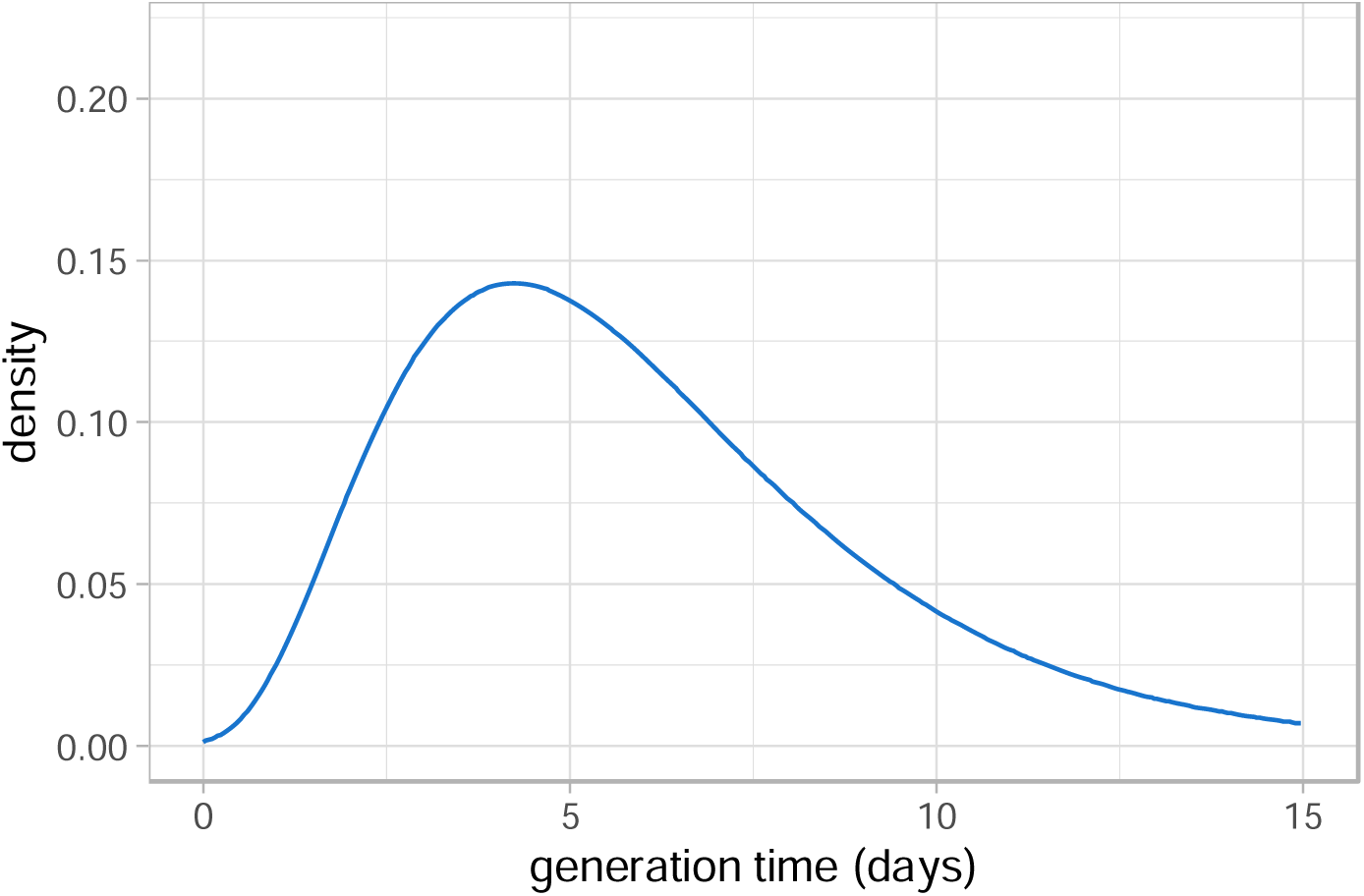
Prior distribution on the generation time implied by the prior distributions on the basic parameters discussed in the Supplementary Technical Appendix. The solid line represents the mean and the shaded region the 95% Credible Intervals.

**Supplementary Figure 8:**
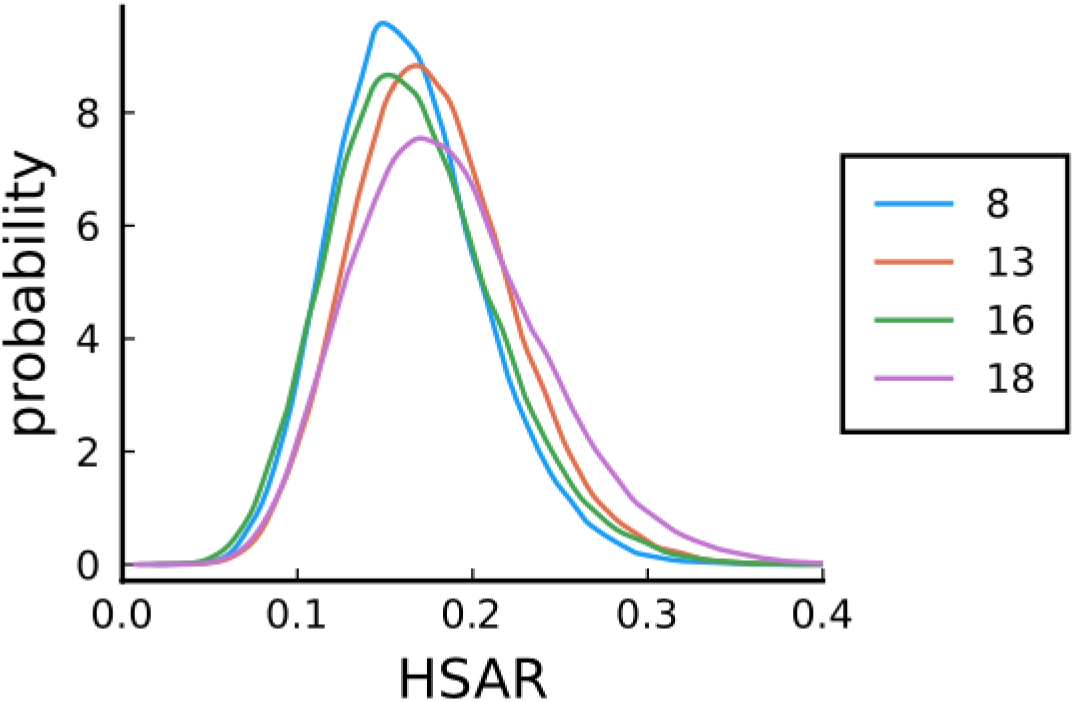
Posterior distributions for the household secondary attack rate using different age cut-offs to define children and adults (in years).

**Supplementary Figure 9:**
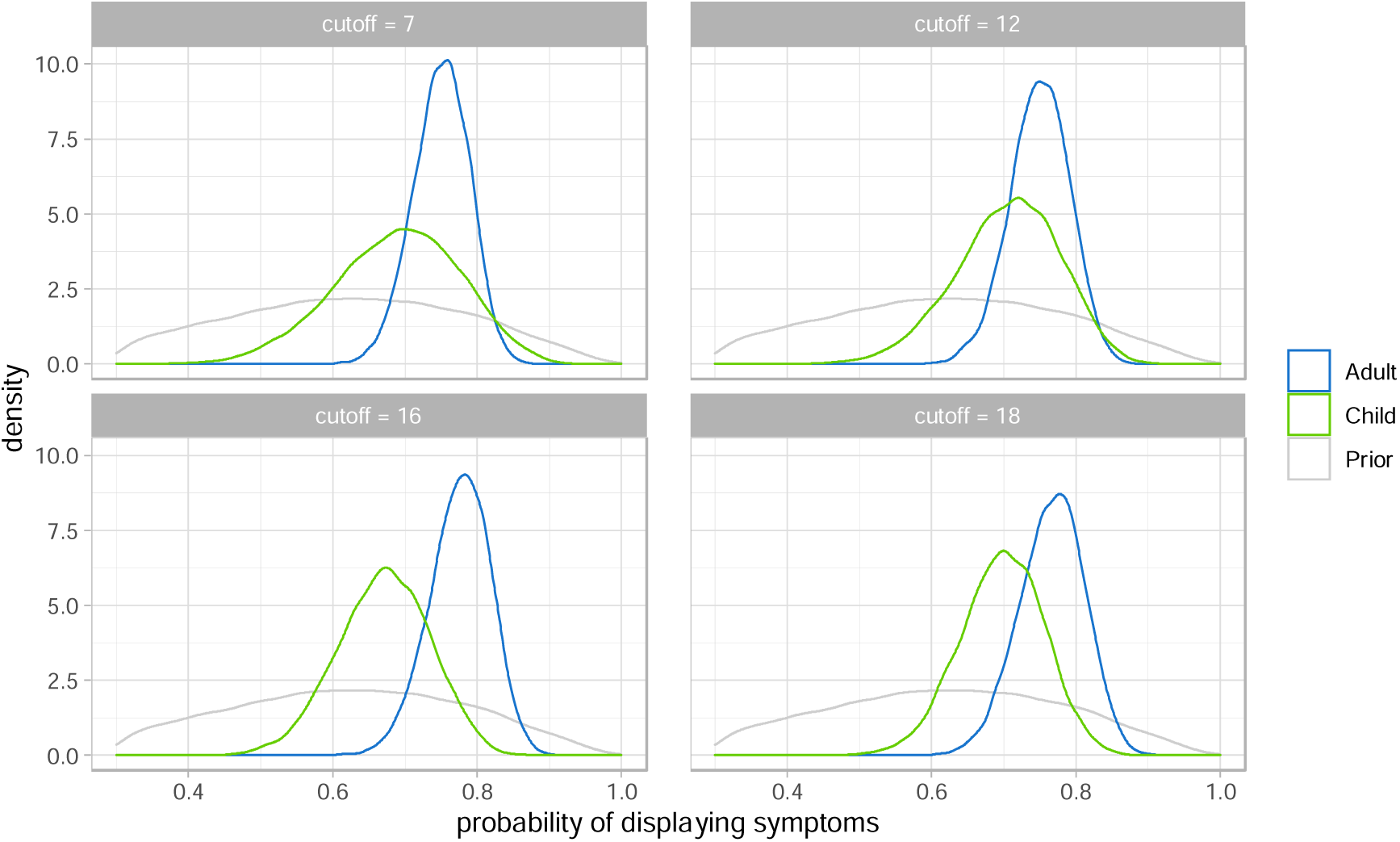
Posterior distributions for the probability of displaying symptoms, conditional on testing positive for adults (blue) and children (green) defined at various age cut-offs (in years). The grey curve indicates the prior distribution.

### Additional model details – modelling appendix

Disease spread within a household was characterised using an SEIR-type compartmental mathematical model previously developed for pandemic influenza^1-3^, adapted to COVID-19. The model allows for pre- and asymptomatic infection status, and is age-structured with age-specific contact rates.^4^ Adults were defined as 18 years old or older, and children were defined as less than 18 years old.

The structure of the model is illustrated in Appendix Figure 1 with associated parameters described in Appendix Table 1. The classes are split into multiple, identical stages, which makes the distribution of time spent within each class Erlang distributed (rather than exponential) to more accurately reflect the distribution time in each period. The number of stages was chosen to reflect evidence about the distributions of the incubation period and/or the generation interval.^5^

The rate of transmission scales depending on the household size, *N*, and the ages of individuals that they make contact within a household. The transmission rate from an infectious adult of age n to a susceptible adult of age m is given by

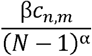

where parameters are as described in Appendix Table 1. The infection rate is scaled based on an individual’s age – specifically, the rate of transmission from an adult to a child is multiplied by *r*_*s*_, the rate of transmission from a child to an adult is multiplied by *r*_*t*_, and the rate of transmission from a child to another child is multiplied by *r*_*s*_*r*_*t*_. The terms *r*_*s*_ and *r*_*t*_ represent the relative susceptibility and transmissibility of children, respectively.

**Appendix Figure 1:**
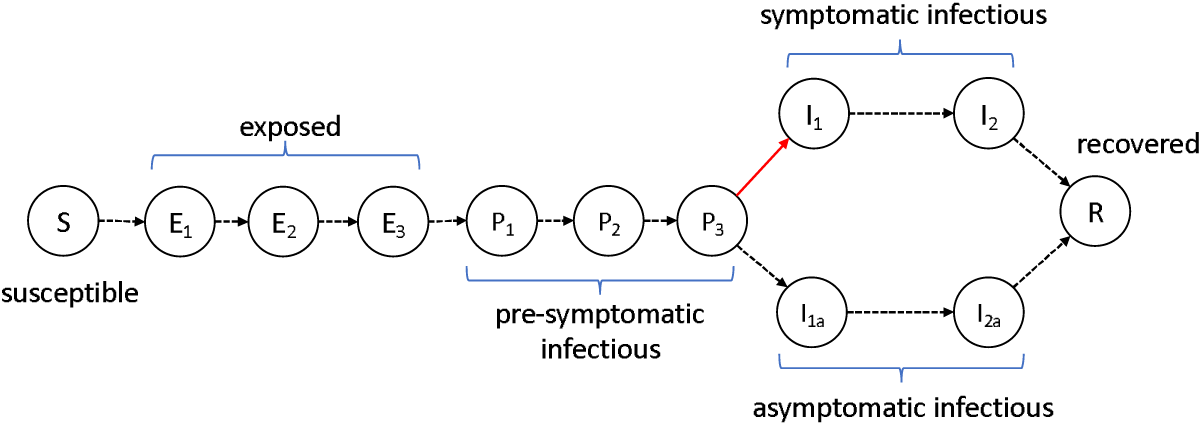
SEIR model structure for the spread of COVID-19 within a household. This illustrates the epidemiological states an individual may be in and how they transition between these states. These states are: S (susceptible); E_1,2,3_ (exposed); P_1,2,3_ (pre-symptomatic and infectious); I_1,2_ (infectious and symptomatic); I_1a,2a_ (infectious and asymptomatic), and; R (recovered). The red arrow indicates the transition when an individual begins to show symptoms. At some point within the infectious period an individual may be hospitalised, and hence removed from the household.

**Appendix Table 1:** Parameters for the model shown in Appendix Figure 1.

| Parameter | Description |
| --- | --- |
| $N$ | Size of household |
| $c_{n,m}$ | Contact rate between age $n$ and $m$ individuals |
| $1/\sigma$ | Average time spent in exposed states |
| $1/\lambda$ | Average time spent in pre-symptomatic states |
| $1/\gamma$ | Average time spent in states $I_1$, $I_2$, $I_{1a}$ and $I_{2a}$ |
| $p_a$ | Probability of developing symptoms for adults |
| $p_c$ | Probability of developing symptoms for children |
| $\beta$ | Rate of infection from adults to adults per contact in a |
|  | household of size 2 |
| $r_s$ | Relative susceptibility of children |
| $r_t$ | Relative transmission from children |
| $\alpha$ | Household size transmission scaling term |

Bayesian inference is performed targeting the parameters of the model using a custom Markov chain Monte Carlo method. The likelihood is estimated using a particle filter that targets the final size of the outbreak within a house, thus we ignore temporal information such as the timing of tests and symptom onset. This approach is adopted as there is very little temporal information and what is available is poorly resolved.

Prior distributions for all parameters of the model are given below:

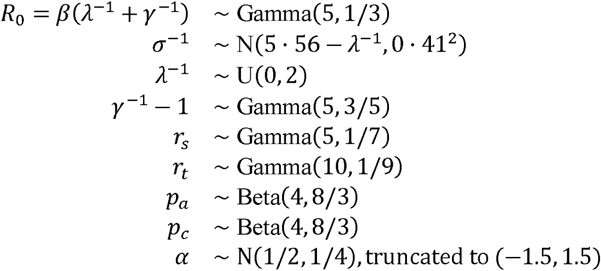

The model priors for the average latent period and pre-symptomatic infectious periods (σ^-1^ and λ^-1^) were fitted using data on exposure windows and symptom onset times from Lauer et al. (2020).^5^ This distribution is calculated using a particle marginal Metropolis Hastings method assuming a uniform [0,10] prior on 1/σ (the average exposed period), a uniform [0,10] prior on 1/λ (the average pre-symptomatic infectious period) and discrete uniform (1,15) shape parameters for each distribution. The resulting joint distribution on *λ*^-1^ and *σ*^-1^ was found to be well approximated by the parametric combination given above.

The prior for the average infectious period γ^*-1*^ was chosen with a mode of 3.4 days and a mean of 4 days. The key quantity which arises from these temporal parameters is the generation time. The distribution for this was obtained by sampling from the joint prior of the parameters (*R*_*0*_, *λ*^-1^, *σ*^-1^, *γ*^*-1*^) and using simulation of the full compartmental model in a fully susceptible population. The final size depends on the transmissibility and generation time distribution, and the effective prior for the generation time distribution is shown in Supplementary Figure 7.^6^ This is consistent with the distribution reported in *Ferretti et. al (2020)*.^7^

Prior distributions were set on the relative susceptibility, *r*_*s*_ and transmissibility, *r*_*t*_ such that the prior distribution for *r*_*s*_ had mode 0.5 and the prior distribution for *r*_*t*_ has mode 1. Each of these distributions are relatively uninformative which captures the prior uncertainty in the parameter values surrounding the differences between children and adults in relation to transmission. The prior distribution on both the observation probabilities was centered around 0.6. The prior distribution on the effect of household size, *α* was taken to be a Normal distribution with mean 0.5 and variance 0.25 truncated on the interval (−1.5, 1.5). This facilitates the possibility of density dependent transmission when *α = 0* and frequency dependent transmission when *α = 1*.

